# External Validation of a Machine Learning Model to Predict Postpartum Hemorrhage in a US Northeastern Healthcare System

**DOI:** 10.1101/2025.08.20.25333642

**Authors:** Vesela P. Kovacheva, Ricardo Kleinlein, Nolan Wheeler, Kartik K. Venkatesh, Eric Jelovsek, David W. Bates, Kathryn J. Gray

**Author notes:** Corresponding Author: Vesela Kovacheva MD PhD 75 Francis St, L1, Boston, MA 02115. These authors contributed equally (Vesela P. Kovacheva, Ricardo Kleinlein). **Twitter (X) post:** Check out our new paper, highlighting that even promising AI models predicting bleeding after childbirth lose accuracy when validated externally. It emphasizes the need for local adaptation and ongoing surveillance in maternal care. Read more: [link] #MaternalHealth #PPH #MachineLearning #EHR #DataScience.

## Abstract

**Introduction:** Postpartum hemorrhage (PPH) is a major cause of maternal morbidity and mortality. Timely prediction may prevent adverse maternal outcomes, and efforts are needed to develop accurate predictive tools. A high-performing machine learning model to predict PPH using data from the US Consortium for Safe Labor (CSL) remains to be widely validated in contemporary clinical settings using electronic health record (EHR) data. Our goal was to evaluate the performance of the CSL PPH predictive model using EHR data across a large healthcare system in the Northeastern US.

**Methods:** We conducted a retrospective cohort study across eight hospitals in the Northeast US between 05/2015 and 05/2024. We used the same sociodemographic, clinical diagnoses, family history, laboratory, and vital signs available on labor and delivery admission in the EHR that were used to train the original CSL model. The binary outcome was PPH, defined as estimated blood loss of 1000 ml or more at delivery or blood transfusion within 24 hours postpartum. We then refit a new model using the original features to assess whether model performance could be further improved in our study population using the best-performing machine learning approach (XGBoost) from the original CSL model. We evaluated model discrimination as measured using the area under the curve (AUC), feature importance, calibration, and decision analysis curves of both the original CSL model with external validation and the further refit model.

**Results:** Among 87,662 deliveries, the incidence of PPH was 7.7%. The original CSL model demonstrated modest discrimination for predicting PPH with an AUC of 0.60 (95% CI, 0.58– 0.61). Refitting a new model with XGBoost resulted in improved discrimination with an AUC of 0.75 (95% CI, 0.74–0.76). Calibration analyses demonstrated that the refit model overestimated PPH risk across a range of predicted probabilities.

**Conclusion:** A promising PPH predictive model had substantially reduced performance with external validation using contemporary EHR data across an eight-hospital health system in the Northeastern US. These findings highlight the importance of external validation, local adaptation, and ongoing surveillance for assessing model performance in an era of evolving prevention, management, and treatment strategies for PPH.

**Key Points:**

- This study aimed to externally validate a previously published machine learning model for predicting postpartum hemorrhage (PPH) and assess its portability across eight hospitals using electronic health record data on labor and delivery admission.
- We found that the original model demonstrated modest discrimination (area under the curve, AUC: 0.60) with external validation.
- A refit model achieved improved discrimination (AUC: 0.75) but remained poorly calibrated and overestimated the risk of PPH across a range of predicted probabilities.
- These findings underscore the importance of local validation and adaptation of external models, and ongoing performance monitoring before clinical deployment of PPH prediction models in an era of evolving prevention, management, and treatment strategies for PPH.

## INTRODUCTION

Postpartum hemorrhage (PPH) is defined as estimated blood loss of at least 1000 mL or blood transfusion within the first 24 hours following delivery,(1) and represents a leading cause of severe maternal morbidity and mortality.(2) The frequency of PPH in the US increased from 2.7 to 4.3% during 2000-2019, which was associated with an increase in the proportion of deliveries with at least one PPH risk factor from 18.6 to 26.9%.(3) Early recognition of PPH, which can result in severe maternal morbidity and mortality, may be facilitated by prompt identification of individuals at risk and timely resource mobilization.(4)

The most widely used tool to predict PPH, recommended by the American College of Obstetricians and Gynecologists and the California Maternal Quality Care Collaborative, is based on expert opinion and often misclassifies risk.(1) For example, only 7% of those classified as ‘high risk’ develop PPH, while 40% who experience PPH were initially considered ‘low risk’.(5) These data illustrate the challenges in accurately identifying PPH risk and have led to the development of multiple alternative tools, such as data-driven predictive models.(6,7) Incorporating a high-performing tool into the EHR could lead to correctly identifying the most at-risk individuals for PPH.

Recent advancements in predictive analytics have generated increasing interest in predicting adverse pregnancy outcomes using EHR.(8,9) Machine learning methods, especially ensemble models, can analyze many complex clinical features simultaneously and can outperform traditional risk scores and regression techniques.(8,9) A promising machine learning PPH model was proposed using data from the US Consortium for Safe Labor (CSL), which included deliveries from >152,000 individuals across 19 clinical sites in the US from 2002 to 2008.(6,10) This machine learning model was highly performing and well-calibrated. A subsequent single-center study using contemporary EHR data and quantitative rather than estimated blood loss was unable to replicate the model’s original performance.(11) Thus, a major challenge in implementing advanced machine learning methods has been their generalizability.(6,7) Models trained on data from one cohort nearly always demonstrate lower performance when applied to an external cohort, commonly due to differences in data quality, patient characteristics, or changes in medical practice.(11,12)

The objective of this study was to examine the performance of the original CSL machine learning PPH prediction model on labor and delivery admission using contemporary EHR data across a large healthcare system in the Northeastern US. We hypothesized that the original CSL model would demonstrate lower performance in our cohort compared with the original CSL dataset and that model refitting would improve its predictive performance.

## MATERIALS AND METHODS

### Study setting and population

We leveraged electronic health record (EHR) data from eight hospitals, two tertiary academic centers, and six community hospitals from the Mass General Brigham (MGB), a large healthcare system in the New England region. We included deliveries between May 31, 2015 (when our institution implemented single-vendor EHR across all outpatient offices and inpatient sites) and May 31, 2024. All data were processed using our machine-learning platform,(13) which extracts and harmonizes data from the EHR. All demographic, family history, medical history, laboratory studies, medications, vital signs, and procedural information data were extracted. Patient race and ethnicity as social constructs were self-reported. We identified the features used in the original manuscript and mapped them in our data. The definitions and sources of each feature are listed in Supplementary Table S1. This study was approved by the MGB’s Institutional Review Board, protocol # 2020P002859, with a waiver of patient consent. We used the TRIPOD reporting guidelines.(14)

Deliveries were included based on documented pregnancies greater than 20 weeks gestation with billing codes for cesarean or vaginal delivery, analyzing each delivery independently.(15) We excluded deliveries with missing blood loss data or >30% missing features.

### Study outcome

PPH was defined as estimated blood loss ≥1000 ml or blood transfusion within 24 hours postpartum to match the CSL model methodology. Due to >10% missing structured blood loss entries, we supplemented data by extracting blood loss information from operative and delivery notes using regular expressions.(16) Deliveries missing documented blood loss (n=1,490) were manually reviewed and excluded.

### Predictive model validation

First, we externally validated the original CSL model using all our study sites. We imputed the mean of the missing numerical features and absence for the missing binary features to remain consistent with the original model methodology. We identified the original 55 features on which the CSL model was trained and followed the methodology of the original study closely.(6)

Next, we refit the original model using the highest-performing machine learning approach used for the original CSL model (XGBoost) in data from all MGB hospital sites, as the new model was intended to be used across all hospital sites. We trained a new XGBoost model using 65% of the MGB data (development) and subsequently evaluated the model performance in the remaining 35% (test). We employed the same set of hyperparameters the pre-trained model was developed with, employing a 10-fold cross-validation approach for the model to only determine in each fold the number of estimators considered. During each fold, we applied early stopping after 100 iterations without improvement on fold-wise validation data, with a maximum of 1,000 estimators, keeping exclusively the best-performing model checkpoint in terms of AUC. To train the final model, we fixed the number of estimators to that of the model in which the AUC score was the highest.

We analyzed the feature importance to better characterize the source of the differences observed between the models using gain visualization and SHapley Additive exPlanations (SHAP) value estimation. Whereas gain represents the improvement in accuracy contributed by a feature to the branches it influences in the model, serving as a straightforward measure of the contribution of a feature during training, SHAP values provide a comprehensive estimation of feature importance by considering the average contribution of each feature across all possible coalitions of features.(17)

### Statistical analysis

Categorical variables were reported as counts and proportions, and continuous variables were reported with medians and interquartile ranges. The Area Under the Curve (AUC) was defined as the ability of the model to discriminate between the positive and negative classes by plotting the sensitivity (true positive rate) and 1-specificity (false positive rate). Bootstrapping was applied to evaluate the statistical significance of differences in model performance. We evaluated model performance using metrics such as accuracy, precision, recall, F1-score, and AUC. Additionally, we calculated calibration and decision curves to evaluate how well these models would perform in clinical practice. We fitted a Platt scaling regressor or a non-parametric isotonic regressor to improve the calibration of our refitted model.(18) Python 3.10 and R 4.4 were used to conduct all analyses. Learning curves, results, and interpretability measurements were visualized using Matplotlib (v3.8.4). Model validation, calibration, and comparison were performed with bootstrapping using the scipy (v1.11.1) package. The pre-trained model was evaluated using the R programming language in which it was initially developed. Several R packages were employed: XGBoost (v1.7.7.1) to manage the pre-trained model and pROC (1.18.5), rms (v6.8.2), and CalibrationCurves (v2.0.3) to carry out statistical analyses.

## RESULTS

### Baseline Characteristics and Outcomes

In this cohort of 137,340 pregnancies at MGB from May 2015 to May 2024, 135,850 had documented blood loss using either estimated or quantitative methods at delivery. We included in the study all 87,662 pregnancies in which blood loss was visually estimated to be consistent with the methods used in the development of the original CSL model (Fig. 1).

**Fig 1.**
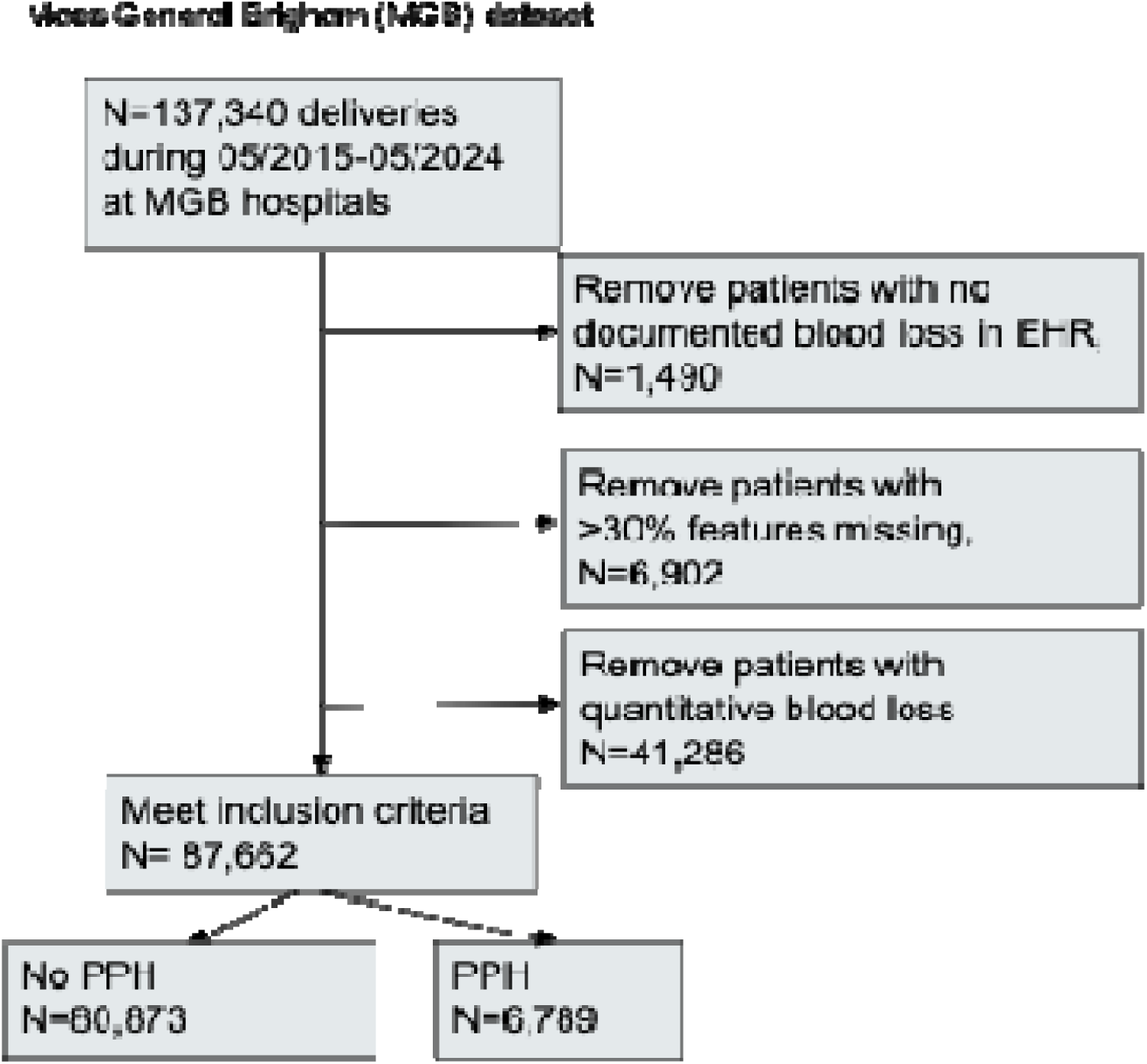
Patient flow diagram. Abbreviations: EHR, electronic health records; PPH, postpartum hemorrhage.

In the included MGB cohort (n=87,662), the mean maternal age was 32.4 years, and the mean BMI was 26.2 kg/m². Self-reported race distribution was 60.3% White (n=52,878), 8.0% Black (n=7,052), 10.0% Asian (n=8,799), and 5.0% identifying as other races (n=4,378).

Additionally, 16.6% of individuals (n=14,555) reported Hispanic ethnicity. Cesarean delivery occurred in 31.9% (n=27,937) of cases. Compared to the individuals in the CSL cohort, those in the MGB cohort were older and had more comorbidities (Table 1).

**Table 1.** Patient characteristics by cohort

| Characteristic | Consortium for Safe Labor<br>(n=228,438) |  | Mass General Brigham<br>(n=87,662) |  |
| --- | --- | --- | --- | --- |
|  |  | Missing |  | Missing |
| Maternal age at delivery (y) | 27.6 ± 6.2 | 323 (0.1) | 32.4 ± 5.07 | 0 (0.0) |
| Self-reported race and ethnicity |  |  |  |  |
| White | 113,224 (51.7) | 9,361 (4.0) | 52,878 (60.3) | 0 (0.0) |
| Black | 51,392 (23.5) | 9,361 (4.0) | 7,052 (8.0) | 0 (0.0) |
| Asian | 9,345 (4.3) | 9,361 (4.0) | 8,799 (10.0) | 0 (0.0) |
| Other | 5,400 (2.5) | 9,361 (4.0) | 4,378 (5.0) | 0 (0.0) |
| Hispanic | 39,716 (18.1) | 9,361 (4.0) | 14,555 (16.6) | 0 (0.0) |
| Academic hospital | 130,205 (57.0) | 0 (0.0) | 59,396 (67.8) | 0 (0.0) |
| Pre-pregnancy BMI (kg/m<sup>2</sup>) | 25.4 ± 6.25 | 76,818 (33.6) | 26.2 ± 5.99 | 26,046 (29.7) |
| Pre-pregnancy weight (kg) | 68.0 ± 17.6 | 67,294 (29.4) | 70.0 ± 16.6 | 23,576 (26.9) |
| Admission weight (kg) | 82.5 ± 17.9 | 35,984(15.7) | 81.7 ± 16.9 | 934 (1.1) |
| Maternal temperature (F) | 98.0±0.81 | 47,977 (21.0) | 99.1 ± 0.85 | 167 (0.2) |
| Systolic blood pressure (mmHg) | 124±14.9 | 52,766 (23) | 139 ± 17.1 | 811 (0.9) |
| Diastolic blood pressure (mmHg) | 74±11.6 | 52,800 (23.1) | 86.2 ± 12.0 | 811 (0.9) |
| Tobacco use | 15,247 (6.6) | 0 (0.0) | 181 (0.2) | 0 (0.0) |
| Illicit drug use | 4,725 (2.0) | 22,544 (9.8) | 1785 (2.0) | 0 (0.0) |
| Parity | Parity 1 (0, 2) | 0 (0.0) | 1.00 [1.00, 2.00] | 33325 (38.0) |
| Gestational age at delivery (weeks) | 39 (38,40) | 0 (0.0) | 38.9 (2.12) | 0 (0.0) |
| Anemia | 23,057(10.4) | 7,877 (3.4) | 12,671 (14.5) | 0 (0.0) |
| Assisted reproductive technology | 1,101 (0.9) | 107,479 (47.0) | 10,197 (11.6) | 173 (0.2) |
| Multiple gestation | 5,053 (2.2) | 0 (0.0) | 2,821 (3.2) | 0 (0.0) |
| Breech presentation | 11,647 (5.1) | 0 (0.0) | 4,946 (5.6) | 0 (0.0) |
| History of preterm birth | 17,017 (7.6) | 5,436 (2.3) | 1,252 (1.4) | 0 (0.0) |
| Polyhydramnios | 885 (0.4) | 14,169 (6.2) | 2,318 (2.6) | 173 (0.2) |
| Fetal macrosomia | 1,858 (1.5) | 102,247 (44.7) | 7,431 (8.5) | 807 (0.9) |
| Cesarean delivery | 65,990 (28.8) | 0 (0.0) | 27,937 (31.9) | 532 (0.6) |
| Prior cesarean delivery | 31,321 (14.5) | 13,219 (5.7) | 1,626 (1.9) | 32,772 (37.4) |
| Spontaneous labor | 122,673 (53.7) | 0 (0.0) | 15,064 (17.2) | 0 (0.0) |
| Trial of labor | 192,074 (84) | 0 (0.0) | 76,359 (87.1) | 532 (0.6) |
| Placenta previa | 1,647 (0.7) | 0 (0.0) | 3,124 (3.6) | 0 (0.0) |
| Placental abruption | 3,794 (1.6) | 0 (0.0) | 473 (0.5) | 0 (0.0) |
| Placenta accreta | 138 (0.0) | 82,335 (36.0) | 7,056 (8.0) | 0 (0.0) |
| Fetal growth restriction | 2,678 (1.2) | 15,626 (6.8) | 4,904 (5.6) | 173 (0.2) |
| Large for gestational age | 2,031 (1.4) | 84,338 (36.9) | 1,474 (1.7) | 173 (0.2) |
| Antenatal steroids | 5,945 (4.0) | 82,507 (36.1) | 3,634 (4.1) | 13,647 (15.6) |
| Antepartum bleeding in third trimester | 3,088 (1.7) | 49,410 (21.6) | 15,427 (17.6) | 173 (0.2) |
| Threatened preterm birth antepartum | 7,880 (3.4) | 0 (0.0) | 2,794 (3.2) | 173 (0.2) |
| Premature rupture of membranes | 16,219 (7.1) | 0 (0.0) | 1,158 (1.3) | 0 (0.0) |
| Chorioamnionitis on admission | 1,527 (1.0) | 83,009 (36.3) | 41 (0.0) | 0 (0.0) |
| Antepartum hospital admission | 17,500 (7.6) | 84,603 (37.0) | 17,484 (19.9) | 0 (0.0) |
| Gestational diabetes | 11,999 (5.2) | 0 (0.0) | 12,269 (14.0) | 0 (0.0) |
| Gestational hypertension | 6,286 (2.7) | 0 (0.0) | 6,022 (6.9) | 173 (0.2) |
| Preeclampsia without severe features | 7,457 (3.2) | 0 (0.0) | 2,020 (2.3) | 173 (0.2) |
| Preeclampsia with severe features | 3,772 (1.6) | 0 (0.0) | 1,236 (1.4) | 0 (0.0) |
| Superimposed preeclampsia | 1,971 (0.8) | 0 (0.0) | 129 (0.1) | 173 (0.2) |
| Eclampsia | 326 (0.1) | 0 (0.0) | 19 (0.0) | 173 (0.2) |
| Fetal death | 1,145 (0.5) | 0 (0.0) | 263 (0.3) | 0 (0.0) |
| Group B Streptococcus colonization | 40,207 (17.6) | 0 (0.0) | 6,856 (7.8) | 0 (0.0) |
| Intrapartum magnesium sulfate | 8,763 (4.3) | 26,136 (11.4) | 4,101 (4.7) | 0 (0.0) |
| Pregestational diabetes | 5,305 (2.3) | 0 (0.0) | 2,446 (2.8) | 0 (0.0) |
| Chronic hypertension | 7,690 (3.3) | 0 (0.0) | 4,391 (5.0) | 0 (0.0) |
| Heart disease | 3,481 (1.5) | 0 (0.0) | 1,837 (2.1) | 173 (0.2) |
| Thyroid disease | 5,583 (2.5) | 7,878 (3.4) | 8,682 (9.9) | 173 (0.2) |
| Renal disease | 1,856 (0.8) | 0 (0.0) | 1,574 (1.8) | 0 (0.0) |
| Asthma | 17,490 (7.6) | 0 (0.0) | 7,059 (8.1) | 0 (0.0) |
| Depression | 9,850 (4.3) | 0 (0.0) | 8,433 (9.6) | 173 (0.2) |
| Gastrointestinal disease | 2,592 (1.1) | 0 (0.0) | 29,156 (33.3) | 0 (0.0) |
| Active seizure disorder | 210 (0.1) | 24,679 (10.8) | 567 (0.6) | 0 (0.0) |
| History of seizure disorder | 1,528 (0.7) | 22,544 (9.8) | 383 (0.4) | 32,772 (37.4) |
| Outcome: postpartum hemorrhage | 7,279 (4.8)<br>(data available for 152,279) | 0 (0.0) | 6,789 (7.7) | 0 (0.0) |

Mean ± standard deviation for continuous variables; n (%) for categorical variables.
Abbreviations: BMI, body mass index.

The rate of PPH, defined as blood loss of at least 1,000 ml in the first 24 hours after delivery, was 7.7% (n=6,789) in the MGB cohort, which was higher than 4.8% (n=7,279) in the CSL cohort. The maternal characteristics of the individuals from MGB based on estimated blood loss are summarized in Supplementary Table S3. Similar to the individuals in the CSL cohort, those in the MGB cohort who developed PPH had higher rates of comorbidities such as preeclampsia, gestational diabetes, and anemia.

### CSL Model Performance in MGB Cohort Data

We evaluated the ability of the original CSL using the machine learning approach of XGBoost to predict the risk of PPH in MGB data. The overall performance in predicting PPH risk, as measured by the area under the curve (AUC), was 0.60 (95% CI, 0.58–0.61), Fig. 2A, which was lower compared with the performance in the original dataset, AUC 0.93 (95% CI, 0.92-0.93). The model also achieved different performance based on hospital and year. Specifically, the model performed slightly better on data by hospital site from Massachusetts General Hospital than on Brigham and Women’s Hospital data, with an AUC of 0.64 (95% CI, 0.63 - 0.65) and 0.56 (95% CI, 0.55 - 0.57), respectively. Additionally, performance improved modestly in deliveries after 2021, with an AUC of 0.61-0.64 (95% CI, 0.58 - 0.67). The model demonstrated slightly lower predictive performance among individuals who self-identified as Black (AUC 0.56; 95% CI, 0.54–0.58) compared with individuals from other self-reported racial and ethnic groups (AUC 0.61–0.62; 95% CI, 0.59–0.65). Additional performance metrics are reported in Supplementary Tables S4-S6.

**Fig 2.**
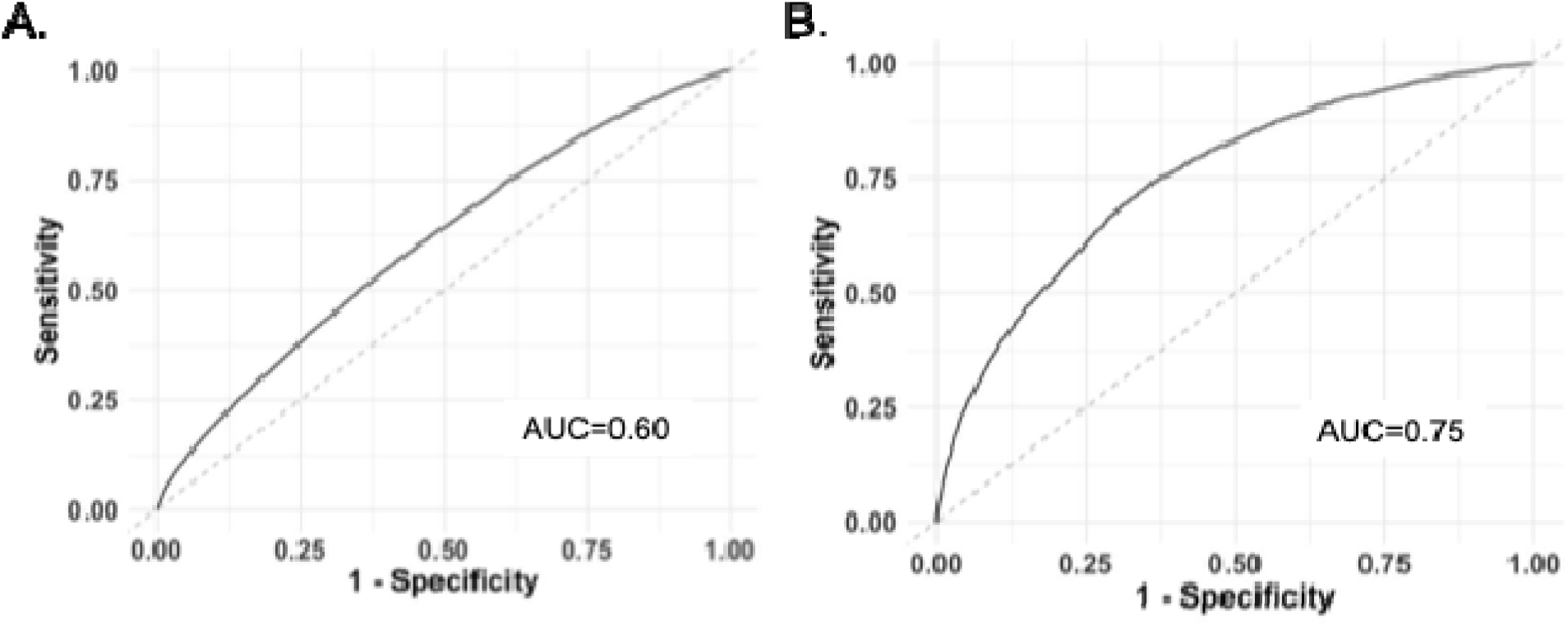
Performance of the models in the Mass General Brigham (MGB) data. (A) Area under the receiver-operator characteristics curve of the original CSL model on the entire MGB data, and (B) the newly refit XGBoost models evaluated on the held-out MGB test dataset.

### Refitting a New Model in MGB Data

We subsequently selected the features used in the CSL model and refitted a new model using MGB data, which led to an improvement in the AUC to 0.75 (95% CI, 0.73–0.76) when considering the prediction over the held-out test set (Fig. 2B). The remaining performance metrics are shown in Table 2.

**Table 2.**
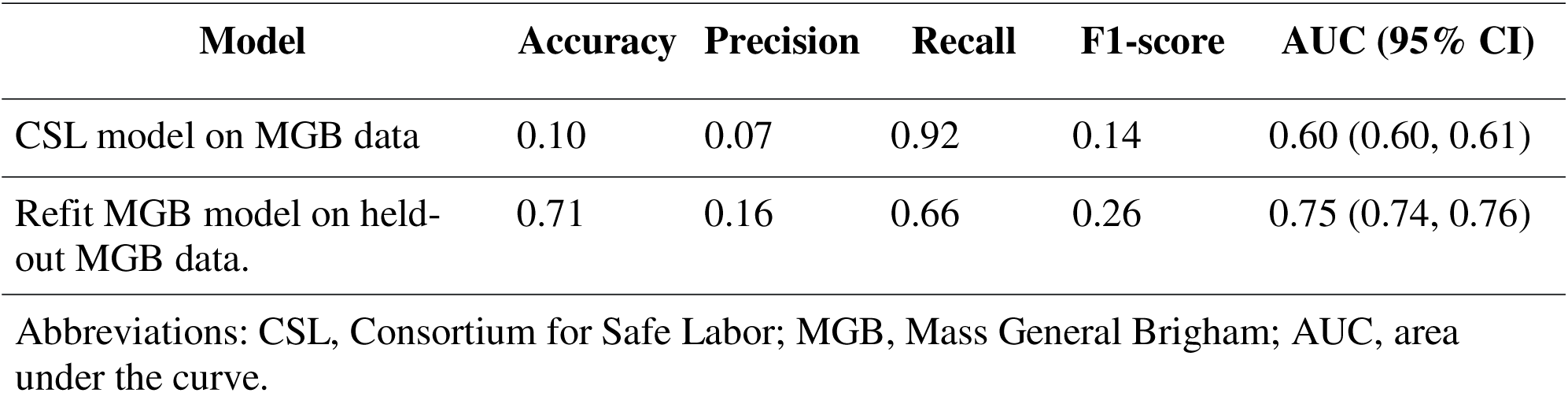
Performance metrics of the CSL model applied to the MGB data and the newly refit model applied on held-out MGB data.

| Model | Accuracy | Precision | Recall | F1-score | AUC (95% CI) |
| --- | --- | --- | --- | --- | --- |
| CSL model on MGB data | 0.10 | 0.07 | 0.92 | 0.14 | 0.60 (0.60, 0.61) |
| Refit MGB model on held-out MGB data. | 0.71 | 0.16 | 0.66 | 0.26 | 0.75 (0.74, 0.76) |

Abbreviations: CSL, Consortium for Safe Labor; MGB, Mass General Brigham; AUC, area under the curve.

Given that models from the extreme gradient boosting family (such as XGBoost) may be prone to overfitting, we also trained a logistic regression model and a Lasso model. However, these models had worse performance than XGBoost across all metrics, such as AUC, accuracy, precision, recall, and F1 score.

### Calibration of the original CSL and refit models

In contrast to the excellent calibration of the CSL model in the original CSL cohort, the model demonstrated poor calibration in the MGB cohort. Among individuals classified as high-risk (defined as having a predicted probability of PPH greater than 80%), the model consistently overestimated the incidence of PPH compared with observed outcomes. Conversely, among low-risk individuals (defined as having a predicted probability below 20%), the model consistently underestimated the actual incidence of PPH (Fig. 3A). The calibration curve for the refit model in the MGB test cohort overestimated PPH risk, demonstrating, that individuals predicted at high PPH risk by the model generally did not develop PPH (Fig. 3B). Post-hoc fitting using either Platt scaling or a non-parametric isotonic regression in our refit model did not improve model calibration.

**Fig. 3.**
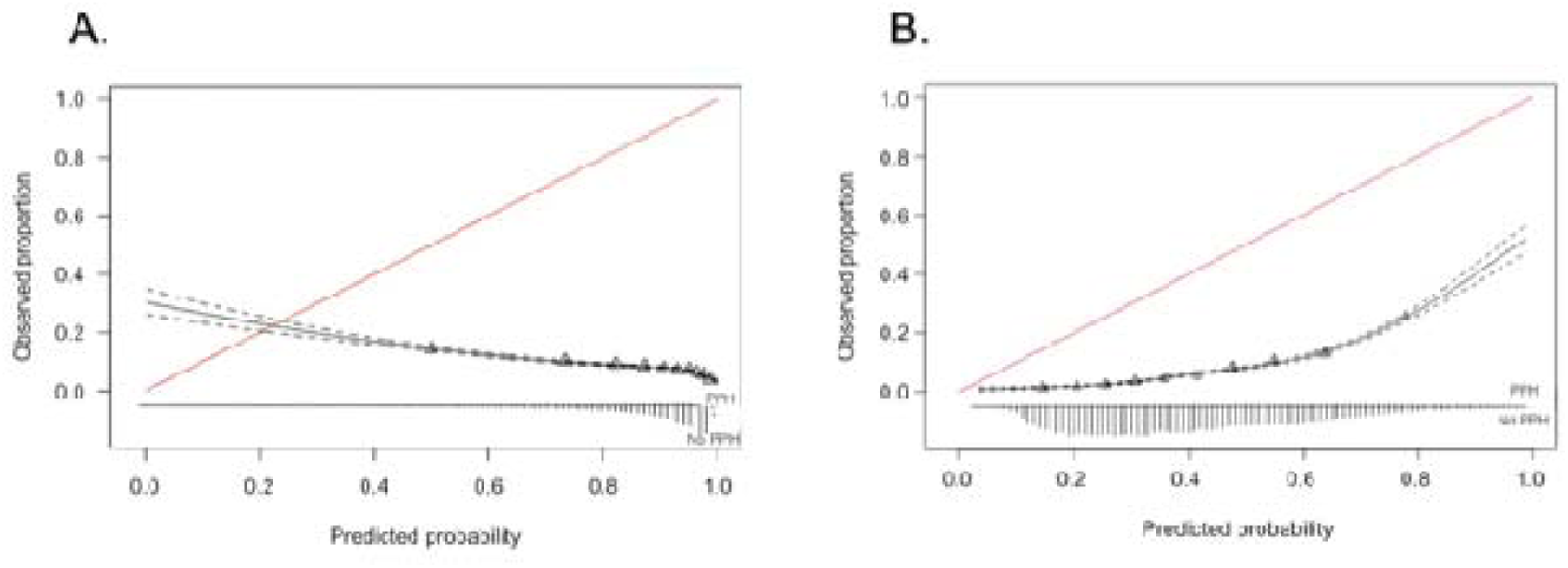
(A) Calibration plot of the original CSL model (held-out Mass General Brigham [MGB] test set); (B) calibration of the locally refit XGBoost model (held-out MGB test set). The red 45° line indicates perfect calibration (predicted = observed). The black line shows the estimated calibration curve; dotted lines indicate 95% CIs. Black triangles are bin-averaged points (deciles of predicted risk).

### Feature Importance

The CSL and the refitted MGB model demonstrated similar importance features such as spontaneous labor, temperature, trial of labor, and weight using gain (Fig. 4A) and Shapley values methods (Fig. 4B).

**Fig. 4:**
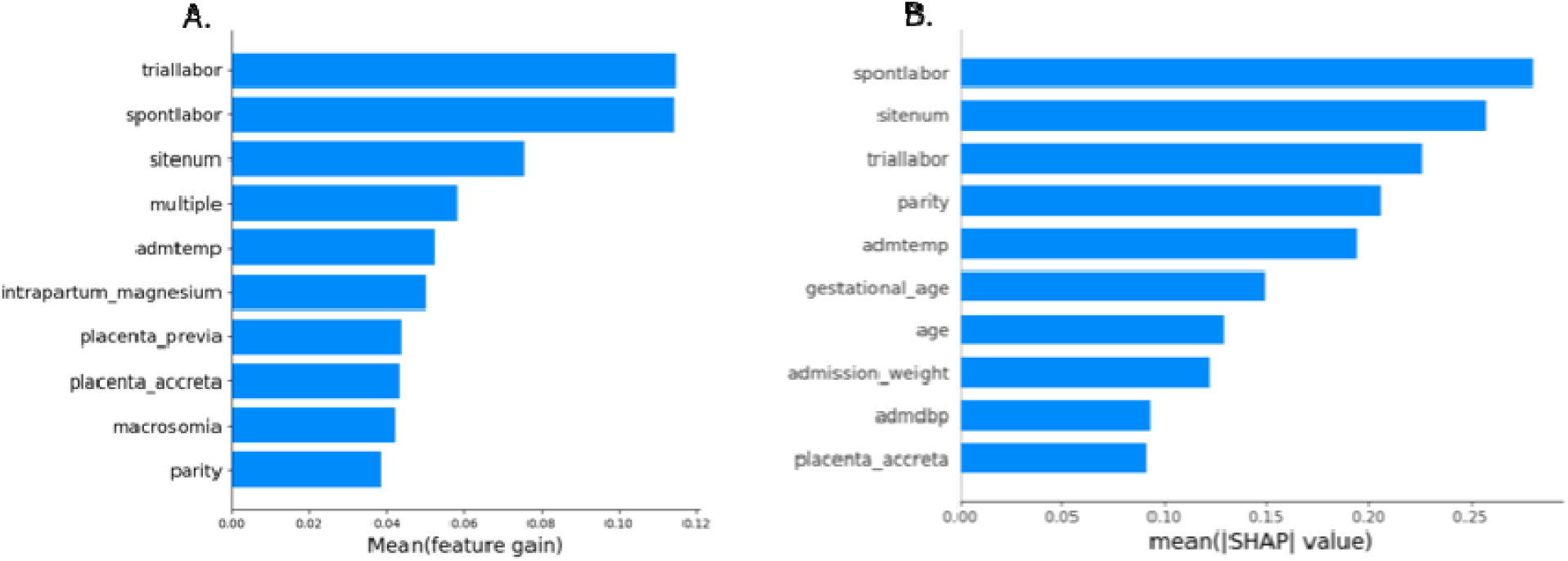
(A) XGBoost model top ten mean feature gain values, and (B) estimated Shapley values for the newly refit XGBoost models in MGB data.

### Clinical Usefulness of the Models using decision-curve analysis

Decision-curve analysis evaluating net benefit at various threshold probabilities showed that both the original CSL model and the locally refit model provided minimal clinical advantage over the strategies of classifying all individuals as low-risk (“treat none”) or as high-risk (“treat all”). Furthermore, the net benefit was very similar between the original CSL and refit models, indicating limited clinical utility for both approaches in the MGB cohort.

## DISCUSSION

We performed an external validation of a recently developed PPH predictive model from the US CSL cohort using contemporary EHR data across eight hospitals in a large Northeastern healthcare system. We found that the CSL model demonstrated substantially reduced discrimination and poor calibration to predict PPH accurately using contemporary EHR data.

Although the CSL model performed extremely well in its original derivation cohort, we observed a marked decrease in performance (AUC of 0.60 in MGB vs. 0.93 originally reported) and inferior calibration when performing external validation. Despite improved model discrimination with refitting (AUC 0.75), the refit model did not result in enhanced clinical utility based on the decision curve analysis. Calibration and net benefit remained low. These results demonstrate the importance of external validation before considering prospective implementation, as the generalizability of models beyond the development cohort cannot be assumed.

Several factors may explain the observed decrease in performance, including differences in patient populations, methods, and evolving clinical practice. Importantly, the CSL data collection protocol was not designed for a machine learning study, potentially allowing inclusion of some features not available at admission.(10) Predictive models frequently demonstrate lower performance in external datasets due to overfitting, bias, and poor reporting quality.(12) To overcome these limitations, we collaborated with the authors of the original model and followed their methodology closely. Similar replication challenges occurred previously, attributed to differences in outcome ascertainment.(11) In one single-center study, the CSL model performed with an AUC of 0.57 on local data.(11) The rate of blood loss was measured using quantitative methods, and the frequency of PPH was higher at 25% compared with 5% in the original CSL cohort and 8% in the current analysis. Implementation of quantitative blood loss measurement methods leads to increased PPH detection and blood transfusion rates.(19,20) Although we excluded quantitative blood loss data, evolving practices at Brigham and Women’s Hospital since 2018 may have introduced variability in data consistency.(21)

In the current study, refitting the model using MGB data led to improved discrimination (AUC of 0.75), suggesting valuable predictive information in the original model features. However, even the locally refit model showed poor calibration, demonstrating systematic bias, similar to others.(11,22) These calibration failures highlight that strong discrimination alone is insufficient; models that are not well calibrated may offer little value for guiding clinical decision-making.

Poor calibration can lead to either over- or undertreatment of individuals. One explanation for calibration failures is that significant improvements in clinical practice over the past decade have altered the relationship between traditional PPH risk factors and outcomes. For example, widespread PPH screening on admission,(23) protocolized management of uterine atony,(24,25) novel use of tranexamic acid,(26,27) and new intrauterine devices have been introduced.(28) As a result, new types of models and additional risk factors may have to be considered.

The net benefit analyses had limited clinical utility, further emphasizing the importance of additional analyses beyond standard performance metrics. Despite improved discrimination, neither the original nor the refitted model demonstrated sufficient net benefit for clinical implementation. Machine learning models for PPH may need to incorporate more granular and dynamic features, such as detailed obstetric management, temporal factors, or improved measures of blood loss, to enhance clinical utility. These considerations extend beyond machine learning models, as logistic regression and rule-based tools for PPH risk have also demonstrated limited clinical utility.(29) The accurate prediction of PPH risk has been fundamentally challenging as unexpected hemorrhage may develop in individuals without known risk factors, and retrospective data may lack sufficient depth for a comprehensive analysis.(29)

Our results highlight an important consideration of predictive model portability in medicine: even high-performing models trained on data from select populations often exhibit reduced predictive accuracy and require local adaptation when applied across different institutions, practice patterns, and patient populations. External validation, while important, does not by itself guarantee readiness for immediate clinical use.(30) Establishing consistent definitions, documentation, and protocolized care should help achieve better patient outcomes,(1) though whether or not this will be sufficient is uncertain. It may be that models should be developed that learn on their own at individual sites or sites within networks. In addition, the integration of predictive analytics tools into clinical workflows requires ongoing recalibration and validation to ensure that predictions remain accurate and clinically relevant. As such, local adaptation, continuous monitoring of model performance, and periodic recalibration should be routine steps in the life cycle of a clinical prediction model.(31)

Future research should focus on creating robust frameworks for ongoing model development, recalibration, and adaptation of predictive models. In particular, incorporating more granular and time-linked features may help close the gap between predicted and observed outcomes. Models need to be tested across different community and academic settings and patient populations to identify the full range of relationships between risk and outcome. Moreover, the dynamic nature of obstetric care—where treatment practices, interventions, and protocols evolve—necessitates models capable of “adapting” over time. Prospective validation studies and pragmatic clinical trials could also generate evidence of whether the real-world use of machine learning tools for PPH can truly improve maternal outcomes.

### Strengths and Limitations

This study was based on a large, multisite cohort that captured a broad and diverse patient population in a real-world clinical practice. Despite its strengths, our study has several limitations. First, we used only the features available in the EHR. Some individuals lacked documented blood loss, and we partially mitigated this by extracting data from clinical notes. Second, we included estimated rather than quantitative blood loss, which may have introduced measurement error. Third, the time frame of data collection and the specific patient population at the MGB may not be representative of all obstetric care settings in other institutions and countries. Future research should explore including additional features, refining blood loss measurement methods, standardizing clinical practice, and extending validation efforts to other practice environments.

## CONCLUSION

The CSL PPH predictive model exhibited substantially reduced predictive performance in contemporary EHR data across an eight-hospital health system in the Northeastern US. Specifically, even after refitting with local data, persistent issues with calibration and limited clinical benefit remained. These findings highlight the importance of external validation, local adaptation, and ongoing surveillance for assessing the performance of such models in light of evolving prevention, management, and treatment strategies. Achieving reliable, generalizable, and clinically meaningful PPH predictive models will require a concerted effort to incorporate accurate data sources, refine modeling methods, establish best practices for continuous model evaluation and updating with evolving obstetric practice, and underscore the importance of evaluating the local performance of such models.

## Data Availability

The data supporting the findings of this study are not publicly available due to privacy and ethical restrictions imposed by the Institutional Review Board (IRB).

## Abbreviations

AUC: Area under the curve
BWH: Brigham and Women’s Hospital
CSL: Consortium for Safe Labor
EHR: Electronic health records
MGB: Mass General Brigham
MGH: Massachusetts General Hospital
NWH: Newton Wellesley Hospital
PPH: Postpartum hemorrhage

