## Supplemental File for "External Validation of a Machine Learning Model to Predict Postpartum Hemorrhage in a US Northeastern Healthcare System"

**Supplementary Online Content**

**External Validation and Local Refitting of a Machine Learning Model to Predict Postpartum Hemorrhage in a Large Multisite Retrospective Cohort**

| **Supplementary Table S1.** Feature definition and data source. | | | |
| --- | --- | --- | --- |
| **Feature** | **ICD 9 code** | **ICD 10 code** | **EHR Table** |
| Maternal age at delivery (y) |  |  | demographics |
| Self-reported race and ethnicity |  |  | demographics |
| Academic hospital |  |  | admissions |
| BMI (kg/m^2^) |  |  | flowsheet |
| Weight (kg) |  |  | flowsheet |
| Maternal temperature (F) |  |  | flowsheet |
| Systolic blood pressure (mmHg) |  |  | flowsheet |
| Diastolic blood pressure (mmHg) |  |  | flowsheet |
| Tobacco use | 649.0 | O99.33 |  |
| Illicit drug use | 648.3, 760.72 | O99.32 |  |
| Parity |  |  | Pregnancy |
| Gestational age at delivery (weeks) |  |  | Pregnancy |
| Anemia | 648.2, 280, 281, 282, 283 | O99.01, O99.02, D50, D55, D56, D57.1, D57.20, D57.3, D57.40, D57.80, D58, D59 |  |
| Assisted reproductive technology | V23.89 | O09.81 |  |
| Multiple gestation | 651, V27.2, V27.3, V27.4, V27.5, V27.6, V27.7 | O30, O31, Z37.2, Z37.3, Z37.4, Z37.5, Z37.6, Z37.7 |  |
| Breech presentation | 652.2, 652.3, 652.8, 652.9 | O32.1, O32.2,  O32.8, O64.1 |  |
| History of preterm birth | 765.21, 765.22, 765.23, 765.24, 765.25, 765.26, 765.27, 765.28 | Z87.51 |  |
| Polyhydramnios | 657 | O40 |  |
| Fetal macrosomia | 656.6 | O36.6 | pregnancy |
| Cesarean delivery |  |  | pregnancy |
| Prior cesarean delivery | 654.2, 669.50, 669.51 | O31.21, O82, V30.01,  O75.82, P03.4, V31.01, V32.01, V33.01, V34.01, V35.01, V37.01, V39.01, Z38.01, Z38.31, Z38.62, Z38.69, Z38.66, O34.21, O34.211, O34.212, O34.219, O75.82, O90.0, O34.219, O34.21, O34.211, O34.212, O34.219, O75.82, O90.0, O34.219 |  |
| Spontaneous labor | 650 | O60.1, O80, O84.0 |  |
| Trial of labor |  |  | pregnancy |
| Placenta previa | 641.01, 641.03, 641.13, 641.11 | O44.03, O44.13, O44.23, O44.33 |  |
| Placental abruption | 641.2 | O45 |  |
| Placenta accreta | 641.1, 641.0 | O43.2 |  |
| Fetal growth restriction | 656.5 | O36.5 |  |
| Large for gestational age | 656.6 | O36.6 |  |
| Antenatal steroids |  |  | pregnancy |
| Antepartum bleeding in third trimester | 640.03, 641 | O20, O46, O44.31 |  |
| Threatened preterm birth antepartum | 644 | O60 |  |
| Premature rupture of membranes | 658.1 | O42 |  |
| Chorioamnionitis on admission | 658.4, 762.7 | O41.1 |  |
| Antepartum hospital admission | 640.03, 641, 644 | O20, O46, O44.31, O60 | admissions |
| Gestational diabetes | 648.8 | O24.4 |  |
| Gestational hypertension | 642.3 | O13 |  |
| Preeclampsia without severe features | 642.4 | O14.0, O14.9 |  |
| Preeclampsia with severe features | 642.5, 642.2 | O14.1, O14.2, O11 |  |
| Superimposed preeclampsia | 642.7 | O11 |  |
| Eclampsia | 642.6 | O15 |  |
| Fetal death |  |  | pregnancy |
| Group B Streptococcus colonization | V02.51, 648.93 | O99.82, B95.1, A49.1 |  |
| Intrapartum magnesium sulfate |  |  | medications |
| Outcome: postpartum hemorrhage |  |  | flowsheet |

| **Supplementary Table S2. Patient counts from each Mass General Brigham hospital** | | |
| --- | --- | --- |
| **Hospital** | **Individuals with blood loss < 1000 ml**  **(n=80,873)** | **Individuals with blood loss >= 1000 ml**  **(n=6,789)** |
| Brigham and Women’s Hospital | 27,249 | 2,578 |
| Massachusetts General Hospital | 26,654 | 2,916 |
| Newton Wellesley Hospital | 16,359 | 782 |
| Salem Hospital | 4,399 | 265 |
| Wentworth-Douglass Hospital | 4,163 | 157 |
| Martha’s Vineyard Hospital | 703 | 17 |
| Nantucket Cottage Hospital | 739 | 29 |
| Cooley Dickinson Hospital | 508 | 38 |
| Unknown/Extramural | 99 | 7 |

| **Supplementary Table S3.** Comparison of patient characteristics in the Mass General Brigham cohort stratified by presence or absence of postpartum hemorrhage. | | | |
| --- | --- | --- | --- |
| **Characteristic** | **Individuals with blood loss < 1000 ml**  **(n=80,873)** | **Individuals with blood loss ≥ 1000 ml**  **(n=6,789)** | **P-value** |
| Maternal age at delivery (y) | 32.4 (5.05) | 33.6 (5.17) | <0.001 |
| Self-reported race and ethnicity |  |  |  |
| White | 49,048 (60.6) | 3,830 (56.4) | <0.001 |
| Black | 6,309 (7.8) | 743 (10.9) | <0.001 |
| Asian | 8,091 (10.0) | 708 (10.4) | <0.001 |
| Other | 4,028 (5.0) | 350 (5.2) | <0.001 |
| Hispanic | 13,397 (16.6) | 1,158 (17.1) | <0.001 |
| Academic hospital | 53,902 (66.7) | 5,494 (80.9) | <0.001 |
| Pre-pregnancy BMI (kg/m^2^) | 26.1 (5.90) | 27.7 (6.76) | <0.001 |
| Pre-pregnancy weight (kg) | 69.7 (16.4) | 73.6 (18.7) | <0.001 |
| Admission weight (kg) | 81.3 (16.6) | 86.5 (18.9) | <0.001 |
| Maternal temperature (F) | 99.1 (0.8) | 99.3 (1.04) | <0.001 |
| Systolic blood pressure (mmHg) | 139 (17.0) | 143 (18.8) | <0.001 |
| Diastolic blood pressure (mmHg) | 86.0 (11.9) | 88.2 (12.9) | <0.001 |
| Tobacco use | 169 (0.2) | 12 (0.2) | 0.854 |
| Illicit drug use | 1,633 (2.0) | 152 (2.2) | 0.469 |
| Parity | 1 [1, 2] | 1 [0, 1] | <0.001 |
| Gestational age at delivery (weeks) | 39.0 (2.05) | 38.3 (2.75) | <0.001 |
| Anemia | 11,298 (14.0) | 1,373 (20.2) | <0.001 |
| Assisted reproductive technology | 9,085 (11.2) | 1,112 (16.4) | <0.001 |
| Multiple gestation | 2130 (2.6) | 691 (10.2) | <0.001 |
| Breech presentation | 4,480 (5.5) | 466 (6.9) | <0.001 |
| History of preterm birth | 1,149 (1.4) | 103 (1.5) | 0.813 |
| Polyhydramnios | 2,020 (2.5) | 298 (4.4) | <0.001 |
| Fetal macrosomia | 6,597 (8.2) | 834 (12.3) | <0.001 |
| Cesarean delivery | 22,362 (27.7) | 5,575 (82.1) | <0.001 |
| Prior cesarean delivery | 1,407 (1.7) | 219 (3.2) | <0.001 |
| Spontaneous labor | 14,605 (18.1) | 459 (6.8) | <0.001 |
| Trial of labor | 71,402 (88.3) | 4957 (73.0) | <0.001 |
| Placenta previa | 2,546 (3.1) | 578 (8.5) | <0.001 |
| Placental abruption | 397 (0.5) | 76 (1.1) | <0.001 |
| Placenta accreta | 6,010 (7.4) | 1,046 (15.4) | <0.001 |
| Fetal growth restriction | 4,561 (5.6) | 343 (5.1) | 0.221 |
| Large for gestational age | 1,303 (1.6) | 171 (2.5) | <0.001 |
| Antenatal steroids | 3,011 (3.7) | 623 (9.2) | <0.001 |
| Antepartum bleeding in third trimester | 13,669 (16.9) | 1,758 (25.9) | <0.001 |
| Threatened preterm birth antepartum | 2,565 (3.2) | 229 (3.4) | 0.667 |
| Premature rupture of membranes | 1,070 (1.3) | 88 (1.3) | 0.983 |
| Chorioamnionitis on admission | 33 (0.0) | 8 (0.1) | 0.0188 |
| Antepartum hospital admission | 15,584 (19.3) | 1,900 (28.0) | <0.001 |
| Gestational diabetes | 11,109 (13.7) | 1,160 (17.1) | <0.001 |
| Gestational hypertension | 5,418 (6.7) | 604 (8.9) | <0.001 |
| Preeclampsia without severe features | 1,764 (2.2) | 256 (3.8) | <0.001 |
| Preeclampsia with severe features | 1,027 (1.3) | 209 (3.1) | <0.001 |
| Superimposed preeclampsia | 113 (0.1) | 16 (0.2) | 0.241 |
| Eclampsia | 16 (0.0) | 3 (0.0) | 0.511 |
| Fetal death | 240 (0.3) | 23 (0.3) | 0.831 |
| Group B Streptococcus colonization | 6275 (7.8) | 581 (8.6) | 0.062 |
| Intrapartum magnesium sulfate | 3405 (4.2) | 696 (10.3) | <0.001 |
| Pregestational diabetes | 2087 (2.6) | 359 (5.3) | <0.001 |
| Chronic hypertension | 3832 (4.7) | 559 (8.2) | <0.001 |
| Heart disease | 1657 (2.0) | 180 (2.7) | 0.013 |
| Thyroid disease | 7869 (9.7) | 813 (12.0) | <0.001 |
| Renal disease | 1408 (1.7) | 166 (2.4) | <0.001 |
| Asthma | 6436 (8.0) | 623 (9.2) | 0.002 |
| Depression | 7646 (9.5) | 787 (11.6) | <0.001 |
| Gastrointestinal disease | 26680 (33.0) | 2476 (36.5) | <0.001 |
| Active seizure disorder | 506 (0.6) | 61 (0.9) | 0.027 |
| History of seizure disorder | 349 (0.4) | 34 (0.5) | 0.224 |
| Mean blood loss (mL) | 393 (231) | 1350 (647) | <0.001 |
| Mean (standard deviation) for continuous variables; n (%) for categorical variables; for *parity,*  median (Q1, Q3); p-values for continuous variables were calculated using the Student’s t-test; for categorical variables based on the Chi-squared test. | | | |

| **Supplementary Table S4.** Classification metrics of the CSL model applied to the MGB data, obtained from different hospitals. | | | | | |
| --- | --- | --- | --- | --- | --- |
| **Hospital** | **Accuracy** | **Precision** | **Recall** | **F1-score** | **AUC (95% CI)** |
| BWH | 0.12 | 0.83 | 0.05 | 0.10 | 0.56 (0.55, 0.57) |
| MGH | 0.11 | 0.75 | 0.02 | 0.05 | 0.64 (0.63, 0.65) |
| NWH | 0.07 | 0.91 | 0.02 | 0.05 | 0.59 (0.57, 0.61) |
| Other | 0.06 | 0.93 | 0.01 | 0.03 | 0.59 (0.57, 0.61) |
| Abbreviations: CSL, Consortium for Safe Labor; MGB, Mass General Brigham; BWH, Brigham and Women’s Hospital; MGH, Massachusetts General Hospital; NWH, Newton Wellesley Hospital; AUC, area under the curve | | | | | |

| **Supplementary Table S5.** Classification metrics of the CSL model applied to the MGB data during different years of delivery. | | | | | |
| --- | --- | --- | --- | --- | --- |
| **Year** | **Accuracy** | **Precision** | **Recall** | **F1-score** | **AUC (95% CI)** |
| 2015 | 0.11 | 0.79 | 0.03 | 0.07 | 0.57 (0.53, 0.62) |
| 2016 | 0.10 | 0.78 | 0.03 | 0.07 | 0.59 (0.57, 0.62) |
| 2017 | 0.11 | 0.86 | 0.04 | 0.08 | 0.60 (0.58, 0.62) |
| 2018 | 0.11 | 0.81 | 0.04 | 0.08 | 0.60 (0.58, 0.62) |
| 2019 | 0.09 | 0.77 | 0.03 | 0.07 | 0.62 (0.60, 0.64) |
| 2020 | 0.10 | 0.84 | 0.03 | 0.05 | 0.59 (0.57, 0.61) |
| 2021 | 0.10 | 0.84 | 0.03 | 0.05 | 0.60 (0.58, 0.63) |
| 2022 | 0.10 | 0.84 | 0.03 | 0.06 | 0.61 (0.58, 0.63) |
| 2023 | 0.09 | 0.82 | 0.03 | 0.05 | 0.62 (0.59, 0.64) |
| 2024 | 0.09 | 0.82 | 0.02 | 0.04 | 0.64 (0.61, 0.67) |
| Abbreviations: AUC, area under the curve | | | | | |

| **Supplementary Table S6.** Classification metrics of the CSL model applied to the MGB data across ethnic/racial groups. | | | | | |
| --- | --- | --- | --- | --- | --- |
| **Hospital** | **Accuracy** | **Precision** | **Recall** | **F1-score** | **AUC (95% CI)** |
| White | 0.09 | 0.81 | 0.02 | 0.04 | 0.61 (0.60, 0.62) |
| Hispanic | 0.10 | 0.81 | 0.03 | 0.06 | 0.61 (0.59, 0.62) |
| Black | 0.16 | 0.83 | 0.07 | 0.13 | 0.56 (0.54, 0.58) |
| Asian | 0.12 | 0.83 | 0.05 | 0.10 | 0.61 (0.59, 0.63) |
| Other | 0.12 | 0.83 | 0.06 | 0.11 | 0.62 (0.59, 0.65) |
| Abbreviations: AUC, area under the curve | | | | | |
